# Health inequities in influenza transmission and surveillance

**DOI:** 10.1101/2020.03.30.20048017

**Authors:** Casey M Zipfel, Shweta Bansal

## Abstract

The lower an individual’s socioeconomic position, the higher their risk of poor health in low-, middle-, and high-income settings alike. As health inequities grow, it is imperative that we develop an empirically-driven mechanistic understanding of the determinants of health disparities, and capture disease burden in at-risk populations to prevent exacerbation of disparities. Past work has been limited in data or scope and has thus fallen short of generalizable insights. Here, we integrate empirical data from observational studies and large-scale healthcare data with models to characterize the dynamics and spatial heterogeneity of health disparities in an infectious disease case study: influenza. We find that variation in social, behavioral, and physiological determinants exacerbates influenza epidemics, and that low socioeconomic status (SES) individuals disproportionately bear the burden of infection. We also identify geographical hotspots of influenza burden in low SES populations, much of which is overlooked in traditional influenza surveillance, and find that these differences are most predicted by variation in susceptibility and access to sickness absenteeism. Our results highlight that the effect of overlapping factors is synergistic and that reducing this intersectionality can significantly reduce inequities. Additionally, health disparities are expressed geographically, as targeting public health efforts spatially may be an efficient use of resources to abate inequities. The association between health and socioeconomic prosperity has a long history in the epidemiological literature; addressing health inequities in respiratory infectious disease burden is an important step towards social justice in public health, and ignoring them promises to pose a serious threat.

**Author summary:** Health inequities, or increased morbidity and mortality due to social factors, have been demonstrated for respiratory-transmitted infectious diseases, most recently evidenced by disparities in COVID-19 severe cases and deaths. Many potential causes of these inequities have been proposed, but they have not been compared, and we do not understand their mechanistic impacts. Our understanding of these issues is further hindered by epidemiological surveillance, which has been shown to overlook areas of low socioeconomic status. Here, we combine mechanistic and statistical modeling with high volume datasets to disentangle the drivers of respiratory transmitted infectious diseases, and to estimate locations where these health inequities are most severe, using influenza as a case study. We show that low socioeconomic individuals disproportionately bear the burden of influenza infection, and that all proposed factors are synergistic in causing these. Thus, public health intervention that targets any one of these drivers may alleviate other issues, as they are not mutually exclusive. Additionally, we provide geographical hotspots for improved surveillance. This work also demonstrates the imperative need to consider inequities and social drivers in data collection, epidemiological modeling, and public health work, as the most vulnerable populations may also be the most likely to be overlooked.

## Introduction

Health disparities are differences in health outcomes between social groups, and they persist in all modern public health settings. Health disparities may be the result of health inequalities, which are caused by biological or cultural variations, or by health inequities, which are driven by unfair factors and are avoidable with policy action [1]. There is extensive evidence that social factors, including education, employment, income, and ethnicity have a distinct influence on how healthy a person is: the lower an individual’s socioeconomic position, the higher their risk of poor health for both chronic and infectious diseases in low-, middle-, and high-income settings alike [2]. There is also a role played by geographic context: the spatial distribution of disparity in health cannot be explained by variation in social factors alone [3]. As the divide in health disparities grows wider across the world and within countries, it is imperative that we continue to understand how social determinants impact health, and how this is reflected geographically [4]. Here, we integrate empirical insights from past studies to characterize the impact of social determinants on the dynamics and spatial heterogeneity in an infectious disease case study, influenza.

Influenza is a respiratory infectious disease that occurs in annual epidemics in temperate regions that can have severe outcomes, especially in young children and elderly individuals [5]. Several studies have demonstrated social differences in influenza morbidity and mortality [6, 7, 8, 9, 10, 11]. For severe influenza, the most impoverished areas have been shown to experience twice the influenza hospitalizations compared to regions with the lowest rates of poverty [12], and low education has been shown to be positively associated with influenza hospitalization rates [13]. Past work has even shown that socioeconomic factors played a significant role in the morbidity and mortality caused by the 1918 influenza pandemic [14, 15, 16]. The proposed determinants of disparities in influenza burden include a number of physiological and socio-behavioral dimensions [17, 18]. In particular, influenza vaccine coverage and healthcare access are higher in areas with increased levels of education and household income [19, 20]. Additionally, low socioeconomic status (SES) individuals have been shown to experience increased susceptibility to respiratory infections due to increased stress [21, 22] and have less access to paid sick leave, resulting in less school and workplace sickness absenteeism, defined as remaining home due to illness [23, 24]. Lastly, it has been proposed that the social patterns of low SES populations affect their influenza risk: larger household sizes and higher population density may lead to higher infection risk [25, 26], while a less robust social network might result in decreased exposure, but also less support during recovery if infected [18].

Mathematical modeling studies of social disparities in influenza burden have used a simulation approach [27, 28, 29] and have focused on the effects of material deprivation (i.e. lack of access from income, education, employment) or social deprivation (i.e. lack of social cohesion and support due to small household sizes, single parenting, divorce or widowing). Such studies are important in uncovering the mechanistic explanations of influenza disparities, but have been limited in their geographical extent, or by the use of proxy measures. For example, [27, 29] consider phenomenological variation in social contact rates without empirical evidence linking vulnerable groups to that variation, thus limiting insights on the mechanisms that lead influenza disparities; [28, 29] focus on dynamics within specific cities, limiting generalizability.

Surveillance-based statistical studies of influenza disparities have been spatial in nature and have highlighted the challenges of disease surveillance under these disparities. Surveillance systems gather the data that shapes our understanding of influenza dynamics, and in the US and most European countries, influenza-like illness (ILI) surveillance occurs through reporting by sentinel healthcare providers. Such sentinel surveillance systems have been resource-efficient means of collecting high quality data, but they do not reliably capture data for all populations, since they are dependent on health care accessibility, health care seeking behavior, and other reporting issues [30, 31]. As a result, studies that rely on healthcare data for characterizing rates of ILI sometimes find decreasing rates of disease with increasing social deprivation [18]. While this negative association may be the result of lower exposure in impoverished areas (as suggested by [18]), it is likely that there exist spatial and social heterogeneities in surveillance caused by healthcare utilization. Indeed, Scarpino et al. have shown that the most impoverished areas are blindspots in the US influenza sentinel surveillance system, ILINet, and models based on these data make the best predictions in affluent areas while making the worst predictions in impoverished locations [32]. To better understand and respond to influenza epidemics and pandemics, we must improve our capability to detect and monitor outbreaks in at-risk populations.

In this work, we (a) develop data-driven epidemiological models to assess how social, behavioral and physiological determinants impact population-level influenza transmission in a controlled manner; and (b) develop statistical ecological models from large-scale disease data to estimate latent influenza burden in vulnerable populations in the United States. We hypothesize that low SES populations bear a disproportionate burden of influenza infection, and that a combination of social, economic and health factors cause this disparity. We aim to identify geographic areas where burden is highest in low SES populations to provide hotspots for additional surveillance. As health disparities widen, it is imperative that we develop an empirically-driven mechanistic understanding of the determinants of health disparities, and capture disease burden in at-risk populations. Such insights can allow for improved influenza forecasting, resource allocation and targeted intervention design.

## Results

Here, we have evaluated the impact of social, behavioral, and physiological mechanisms on driving influenza disparities. We achieved this through epidemiological model experiments in a population network with realistic SES-based contact patterns. This increases our understanding of the role that SES-driven variation plays in determining influenza dynamics. This also allows us to disentangle the effects of multiple proposed drivers of influenza transmission among those of differing SES. We have also assessed the impacts of low SES on influenza at the population level. We estimated low SES ILI incidence rates at the county-level in half of the states in the US, accounting for transmission trends identified in the prior epidemiological model experiments, variation in social, economic and health factors, and measurement biases. This provides estimates of ILI incidence rates among low SES populations at a fine spatial scale, identifying areas which are likely currently overlooked by influenza surveillance systems. These findings also provide an understanding of SES-based factors associated with disproportionate burden at the population level, which could guide future public health efforts to reduce socioeconomic health disparities.

### Contact patterns vary by socioeconomic status

Contact patterns have been demonstrated to vary by socioeconomic status [18], but we have lacked social contact networks that explicitly incorporate these differences. To enable testing of hypotheses about social contact trends, we used an egocentric exponential random graph model (ERGM) to simulate networks with realistic social contact patterns based on socioeconomic status (measured by education level, [33]) from the POLYMOD social contact survey, a large social contact survey conducted across Europe [34] (Additional model details can be found in Methods). The fitted network model is consistent with the contact heterogeneity in the data (Fig 1A), and all individuallevel attributes (i.e. age, sex, contact location, and education level) are significant in predicting contact structure (Table S1). Additionally, we incorporated varying levels of low SES individuals into the networks to investigate hypotheses in populations with varying SES composition (details in Methods). The resulting networks are consistent in network structure based on degree and assortative degree (number of contacts with those of the same attribute) by SES-status (Fig 1B). Thus, networks with increased representation of low SES individuals maintain the same SES-based contact patterns as the POLYMOD data. Importantly, the network model captures variation in contact structure by SES. In particular, low-SES individuals have lower mean degree and variation in degree (Fig 1C), but have higher SES-assortative degree compared to those of higher SES (Fig 1D).

**Figure 1:**
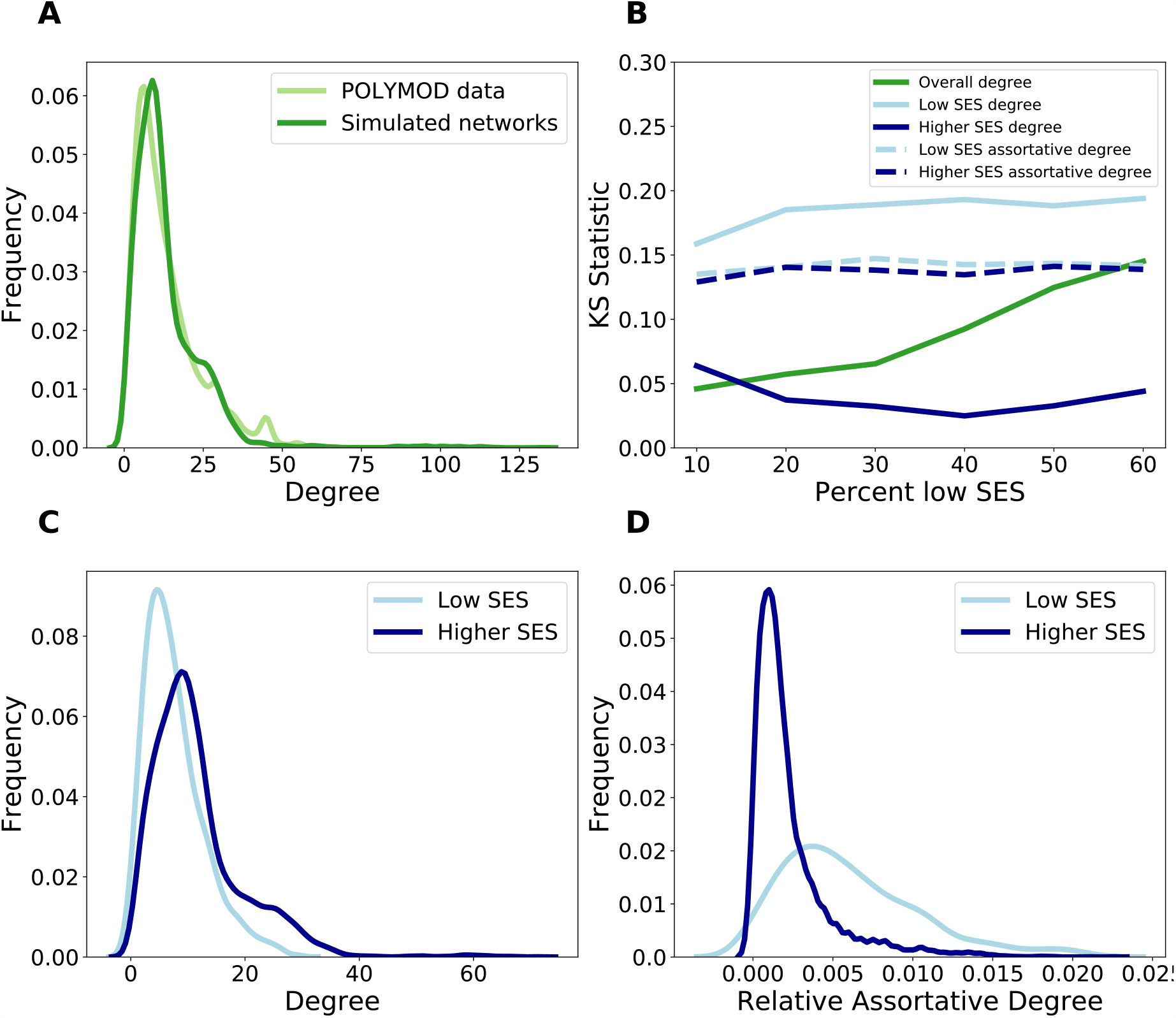
The network characteristics of the networks generated from the ERGM model based on POLYMOD data. A: The degree distribution of the POLYMOD data (light green) compared to the simulated networks (dark green). B: The Kolmogorov-Smirnov (KS) statistic to evaluate the dissimilarity of the ERGM-simulated networks to the POLYMOD data as additional low education individuals are added to the network. KS statistics compare the dissimilarity of the overall degree distribution (dark green), the degree distribution of low SES nodes (light blue, solid), the degree distribution of higher SES nodes (dark blue solid), the assortative degree (e.g. the low SES contacts of low SES nodes) for low SES nodes (light blue, dashed), and the assortative degree for higher SES nodes (dark blue, dashed). Low KS values indicate similar distributions. C: The degree distribution of low SES nodes (light blue) and higher SES nodes (dark blue). D: The relative assortative degree distribution (e.g. number of low SES contacts of low SES nodes/number of low SES nodes) of low SES nodes (light blue) and other SES nodes (dark blue).

### Inequities increase low SES influenza transmission

There appears to be variation in contact trends dependent on socioeconomic status, thus it is important to consider how this network structure impacts epidemiological dynamics. To assess the role of behavioral and physiological heterogeneity, we integrated into an epidemiological network model of influenza transmission five key hypothesized drivers of disparities in influenza burden: a) social contact differences, or fewer social contacts and higher assortativity (as represented in our empirically-informed contact network model); b) low vaccine uptake; c) low healthcare utilization, which results in less access to influenza antivirals; d) high susceptibility, which results from stressful environmental factors; and e) low sickness absenteeism from school or work. Fig 2A shows the infection burden of low SES individuals (i.e. the ratio of the number low SES infections and the number of all infections) in the presence of each factor, combined with social cohesion (included in the network structure). The results are compared against a positive control (light green), in which there is no SES-based heterogeneity in that factor, and mechanisms are randomly distributed throughout the population. Each factor results in a significant increase in the low SES infection burden in the presence of SES-based heterogeneity, and the effect is most pronounced when all the factors occur simultaneously. In contrast, the epidemic size (i.e. the ratio of the number of infections and the population size) for the positive control is larger than the SES-heterogeneous treatments, for all treatments (with the exception of the increased stress treatment) (Fig S32).

**Figure 2:**
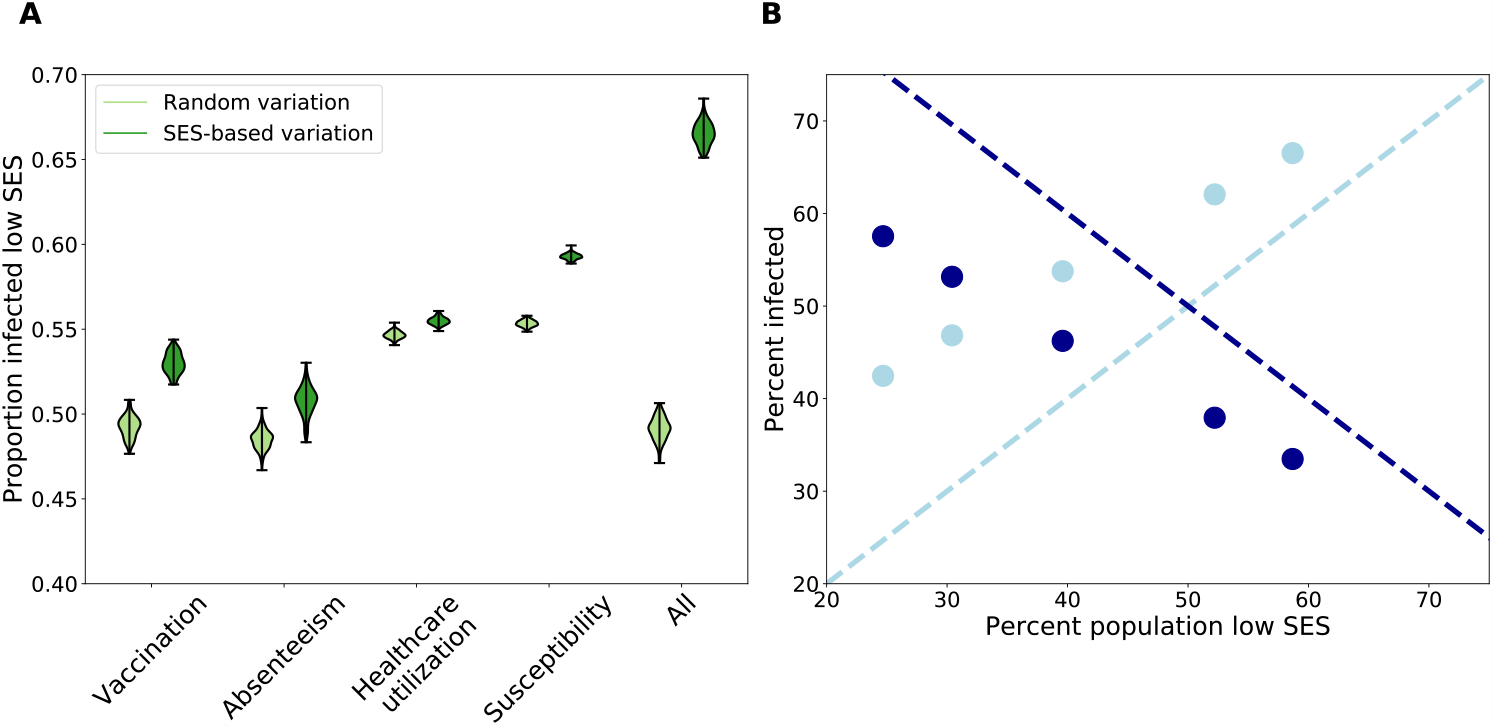
Results of epidemiological simulations on ERGM networks with SES-driven behavioral and physiological differences. A) All of the proposed SES-driven behavioral and physiological differences result in an increase in infection of low SES individuals (dark green, right of paired violin plots), compared to simulations where the differences are randomly distributed throughout the population (light green, left of paired violin plots). This difference is most pronounced when all of the mechanisms occur together. These simulations were performed on a network composed of 60% low SES, but the results are consistent across networks with different SES compositions. B) In all networks, when all SES-driven behavioral and physiological differences are present, low SES individuals (mean percent of infected population that is low SES shown in light blue dots) are disproportionately infected, relative to the expectation (light blue dashed line). High SES individuals are disproportionately underinfected compared the expectation (dark blue dots compared to dark blue dashed line).

This combination of results can be explained by the role that low SES individuals play in the network. On the one hand, low SES individuals have lower mean degree (Fig 1C). When these low degree individuals experience transmission-increasing mechanisms, this results in a smaller epidemic size, compared to the scenario where high SES, and high degree, individuals experience the same mechanisms. Thus, when SES-driven processes that increase transmission affect low SES individuals, it results in a smaller overall epidemic. On the other hand, low SES individuals have high assortativity with other low SES individuals (Fig 1D). Thus, when health disparities increase transmission for low SES individuals, they are more likely to infect other low SES individuals that are also experiencing these mechanisms, resulting in increased spread among this assortative group. This result highlights the need for surveillance and research focused on low SES populations, as the emergent high infection burden of low SES, at-risk individuals could be overlooked due to lower epidemic sizes when aggregated.

Next, we consider how low SES infection burden scales with an increasingly large low SES population. We find that epidemic size increases with an increasing proportion of low SES individuals, and this effect appears to be driven by increasing infection of low SES individuals as they make up a larger component of the network (Fig S31). Indeed, low SES individuals experience a disproportionately large infection burden when all SES-based behavioral and physiological factors occur (Fig 2B). Additionally, high SES individuals experience a disproportionately small infection burden in the presence of the same factors.

### Low SES infection burden is spatially heterogeneous, and high in the southeastern US

Our results thus far characterize the mechanistic role that social, behavioral and physiological factors play on influenza burden in low-SES populations in data-driven controlled experiments. Here, we aim to characterize how macroscopic factors impact influenza dynamics in low-SES populations, integrating our theoretical findings with population-level data. For population-level influenza data, we used medical claims of ILI at the county level in 25 states in the US, based on sufficient data availability. This data stream has been demonstrated to provide enhanced surveillance opportunities for influenza-like illness [31, 35]. However, we find that these data suggest that ILI burden decreases with increasing low SES representation (measured by proportion of low education individuals) (Fig S33). This pattern is counter to our previous mechanistic model findings and to past small scale studies, suggesting that there may be measurement biases in these surveillance data.

To better estimate influenza burden in low SES populations, we fit a Bayesian spatial hierarchical model that accounts for measurement biases and borrows information from spatial covariates pertaining to low SES individuals and the mechanistic modeling experiments (details in Methods). Our model estimates of the low SES ILI incidence ratio (defined as low SES ILI divided by total visits per 1,000 people) show a positive relationship with low SES population size (Fig S38), and allow us to consider spatial disparities in influenza burden. Fig 3 shows the county-level map of the low SES ILI incidence ratio. This map highlights areas with a high incidence rate among low SES individuals in the southeastern United States, which is a region where low SES population levels are high. This also demonstrates that there are significant levels of heterogeneity both within and between states. These estimates can guide targeting of improved surveillance and steps to alleviate the influenza burden in low SES populations.

**Figure 3:**
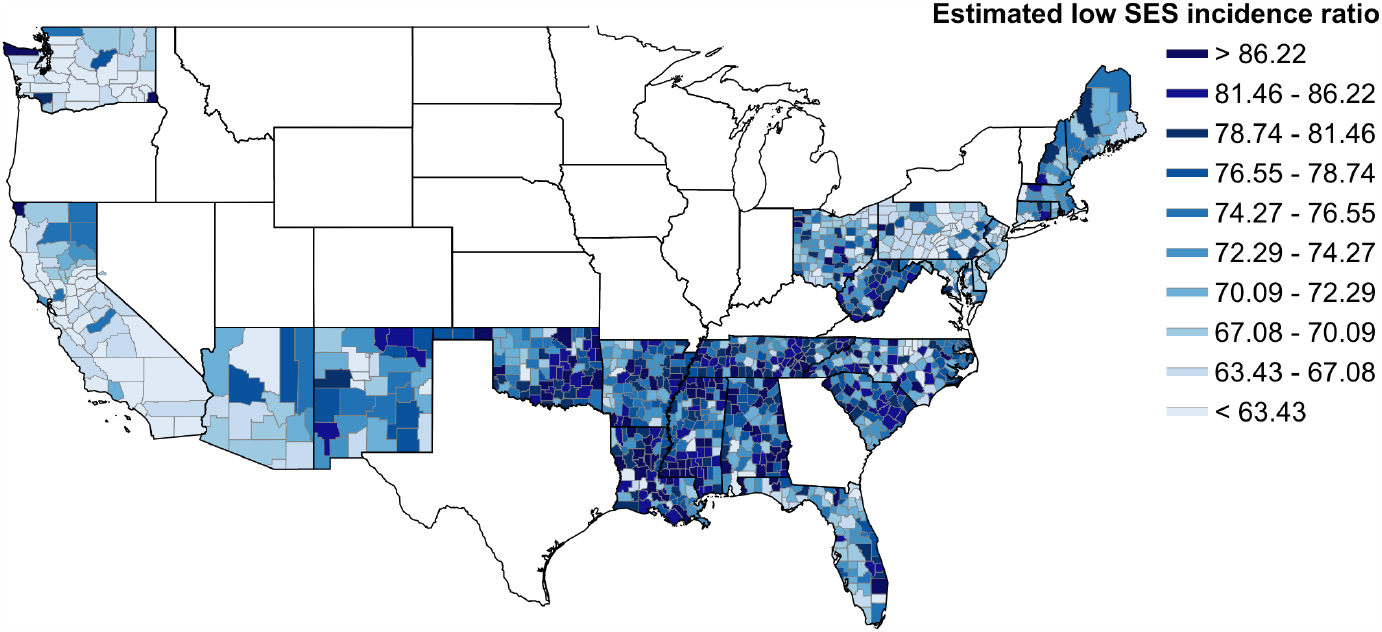
County-level map of model estimates of low SES ILI incidence ratio per 1,000 people. Lower values are represented in light blue, and higher values are represented in darker blue. States in white were omitted due to lack of covariate data.

To validate our findings, we grouped our model estimates by county-level poverty rates, and compared the incidence ratio to prior population-level studies that correlate influenza rates and poverty levels, though these studies do not focus on low SES individuals, so the comparison is not direct. We find increasing low SES ILI incidence rates in areas with increasing levels of poverty, which agrees with trends in [12, 11] (Fig S39). Our results show more consistently high incidence rates compared to the larger increases between poverty rates in prior studies. We attribute this to the incorporation of the measurement process into our models, which accounts for undersurveillance of low SES infection, whereas healthcare access and healthcare seeking differences may have missed low SES cases in prior studies. Ideally, data on respiratory infection of low SES individuals would be available at a fine spatial scale to more directly assess the validity of our models, but the lack of such a dataset highlights the need for future surveillance and data collection that focuses attention on lower SES populations.

### Susceptibility and sickness absenteeism differences are the strongest drivers of ILI in low SES populations

Fig 4 shows the coefficient estimates and credible intervals resulting from the Bayesian spatial hierarchical model. Levels of poor health among low SES individuals, as a measure of susceptibility to infection, are positively related with low SES ILI incidence. Thus, areas with more reports of poor health among low SES individuals exhibit higher incidence rates of low SES ILI. Also, access to sickness absenteeism among low SES individuals, represented by the number of low SES students that are absent for more than 10 days in a school year, is negatively related to low SES ILI incidence rates. Thus, areas where more low SES students are able to be absent experience lower rates of low SES ILI. Student absenteeism may not be a perfect measure of sickness absenteeism or paid sick leave access, but other fine-scale data was lacking, and a student’s ability to be absent is related to a parent’s ability to be home to care for the child, and differences in access to paid sick leave by SES have been related to student sickness absenteeism levels due to influenza [36, 37, 38, 39]. Additionally, while not statistically significant, the model estimates lend evidence for a negative relationship between low SES mean household size and increased low SES influenza incidence; and a positive relationship between low SES influenza vaccination and low SES influenza incidence rates. The household size result provides support to the social deprivation hypothesis that low SES individuals may have less robust social networks, and this may relate to factors that increase transmission, such as increased stress, lack of support when ill that delays recovery, or the need to attend work when ill [18]. The vaccination result is contrary to our expectations for the protective effects of vaccination. This result could reflect vaccination-seeking behavior being higher in areas where influenza rates are typically high thus increasing disease risk perception [40]. Additionally, some prior studies have shown mixed results regarding the relationship between socioeconomic status and influenza vaccination, and that these differing results may relate to the way that socioeconomic status is defined, or what location the study takes place in [41, 42]. Lastly, this may be a result of the source of our vaccination data, which has low coverage in some areas and may suffer from its own measurement biases, which highlights the need for more data on influenza vaccine uptake and the characteristics of those who vaccinate.

**Figure 4:**
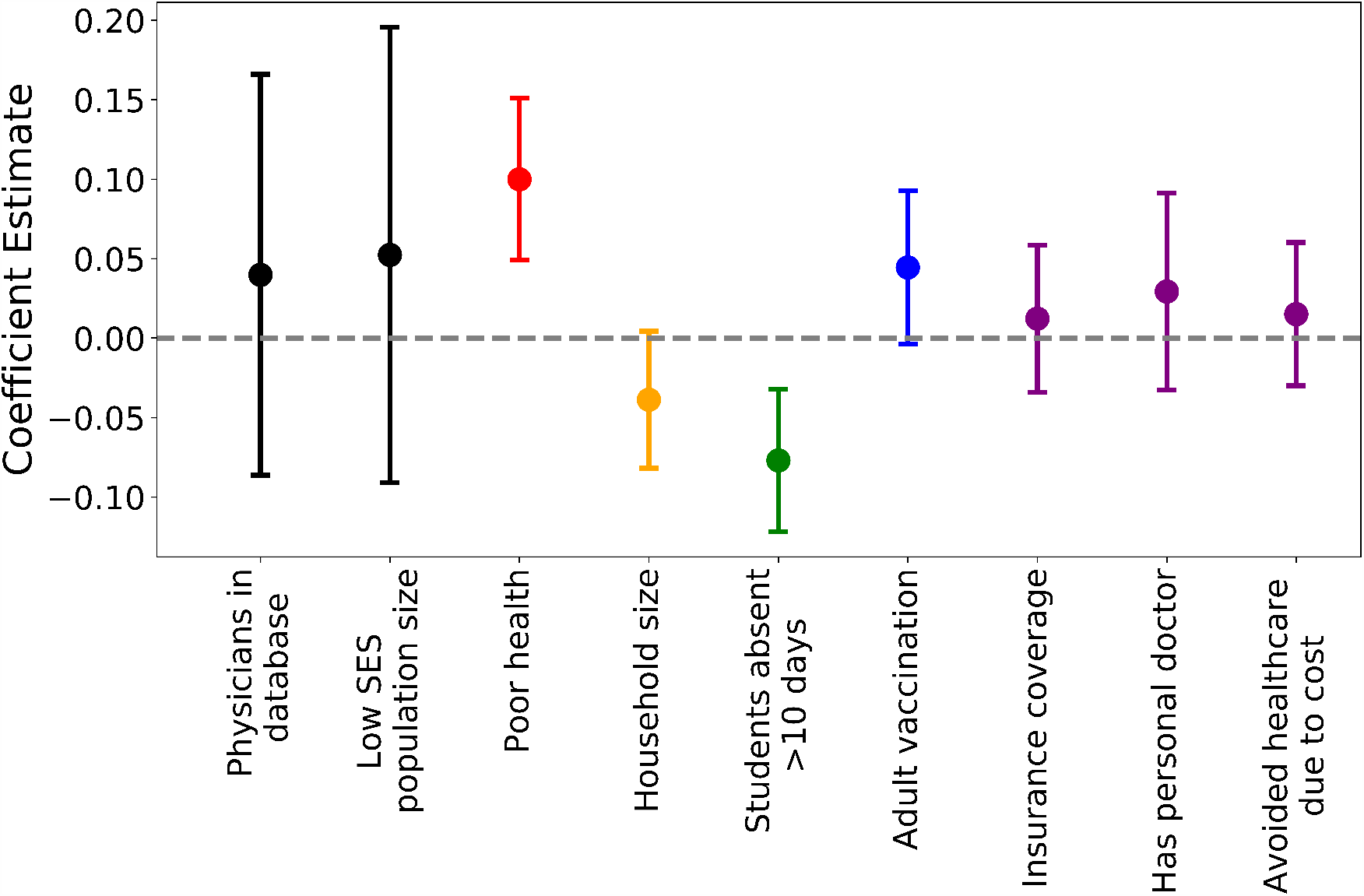
Mean model coefficient estimates and credible intervals. Points are colored by what process each covariate represents (black: measurement bias, red: susceptibility, orange: social contact differences, green: sickness absenteeism, blue: vaccination, purple: healthcare utilization).

## Discussion

Increased infectious disease prevalence among lower socioeconomic status populations has been observed in many settings. What has been missing, however, is a better understanding of the mechanisms that drive this disparity. We used a mechanistic epidemiological network model which allowed us to assess the impacts of SES-based behavioral and physiological differences on influenza in controlled experiments. This highlighted the role played by all mechanisms in tandem to produce disproportionate disease burden in low SES populations. To address the gap that exists in our surveillance of ILI and to estimate the spatial distribution of influenza disparity, we then used a Bayesian spatial hierarchical model to estimate population-level low SES ILI at a fine spatial scale across the United States, accounting for disproportionate infection of low SES individuals, measurement biases, and county-level factors hypothesized to be associated with influenza and SES. Our results shine light on the spatial distribution of respiratory disease health disparities.

In our epidemiological model, disease transmission occurs over the contact network structure, which accounts for heterogeneity in contact patterns by SES. While past work has integrated contact heterogeneity by other socio-demographic characteristics such as age and occupation [43, 44], SES-based contact heterogeneity has not been integrated into contact network models for epidemiological purposes. Epidemiological simulations on the SES-heterogeneous network reveals that each hypothesized behavioral and physiological factor leads to increased infection of low SES individuals. Additionally, we find that communities with larger low SES populations experience larger epidemics, which is in agreement with prior studies [11, 10, 12]. The proposed drivers are not mutually exclusive, so this reveals potential effects that could not be identified in past studies that investigate the impact of a single SES-based mechanism or impacts that might be aggregated in observational studies. We note that these experiments also include SES-based variation in social cohesion (i.e. SES-based contact heterogeneity in the population model), so the effect shown in Fig 2 is the result of both mechanisms combined. In Fig S32, we also illustrate the impact of each mechanism independent of social cohesion.

Our efforts to consider the impacts of low SES on influenza spatial heterogeneity generated county-level maps of ILI incidence in low SES populations. Our findings identify pockets of high ILI burden in low SES populations across the United States, and represent a first step in filling the gap that exists in all healthcare-based surveillance. The model also produced a set of estimates for the effect of each hypothesized ecological measure. We find that low sickness absenteeism and high susceptibility are significantly associated with influenza in low SES populations. This supports our previous finding that multiple mechanisms compound to result in disproportionate low SES influenza burden. To validate our findings, we compared the trends in our model estimates to previous estimates of influenza incidence rates, stratified by poverty level. This is not a direct comparison, as previous studies present the incidence rates for the entire population, not just for low SES individuals within those populations. SES-stratified influenza data would be important to ground truth our model estimates.

Our work has several limitations. The network structure of our epidemiological model is based on one social survey from 2007 in Europe, and may be less representative of the United States today. Additionally, survey data was not collected for the SES of the contacts of survey participants, which required us to make assumptions which could affect our results about SES assortativity. Additional social contact data collection across the United States that accounts for SES heterogeneity would be useful for future studies given the large socio-economic inequality in the country [45, 46]. In our spatial ecological model, we assume that disproportionate burden in low SES populations remains constant over influenza seasons. While this is a reasonable first assumption based on social and healthcare processes being consistent over our study period, there may be variation in the impact of ILI on low SES populations due to strain distribution and environmental features that do vary across seasons. Future work could focus on temporal variation in low SES ILI dynamics. Additionally, our spatial ecological model is only able to provide estimates for half of the states in the US, and the states are mostly on the coasts. This highlights the need for more data collection pertaining to low SES individuals, not only for epidemiological data, but also for a wide variety of other topics to provide covariate data and to create a better understanding of at-risk populations.

As the divide in health disparities grows wider across the United States, we propose the use of infectious disease case studies to improve our understanding of this challenging problem. We suggest that we move beyond studies based on proxy measures such as income and education which may provide an incomplete picture [3], and dig into the mechanisms that may be at the root of inequities. Furthermore, we advocate for the prioritization of capabilities to detect and monitor outbreaks in at-risk populations so that we may prevent exacerbation of health disparities. Addressing health inequities in respiratory infectious disease burden is an important step towards social justice in public health, and ignoring them promises to pose a serious threat to the entire population. Indeed, the damaging impacts of health inequities for respiratory infectious diseases have already been highlighted in the COVID-19 pandemic [47]. Our results suggest that (a) the effect of overlapping behavioral and social factors is synergistic and reducing this intersectionality can significantly reduce inequities; and (b) health disparities are expressed geographically and targeting public health efforts spatially may be an efficient use of resources to abate inequities. Further attention to the mechanisms and processes that lead to health inequities, and specifically health inequities that may be overlooked by our currently surveillance systems, will be important to identifying actionable steps to mitigate negative health outcomes in the future.

## Methods and Materials

In this study, we use (1) a mechanistic network epidemiological model to assess influenza transmission in the presence of individual-level socioeconomic status (SES)-based behavioral and physiological variation; and (2) an inferential spatial model to geographically localize influenza-like illness (ILI) burden among low-SES populations in the presence of population-level variation in social and health indicators. Data and code for the implementation of these methods is available at [48].

### Modeling Impact of Individual-Based SES factors on Disease Burden

To achieve the mechanistic understanding, we (a) fitted a contact network model from empirical contact data that includes contact heterogeneity stratified by age, sex, contact location, and socioeconomic status; and (b) performed epidemiological simulations on these networked populations integrating epidemiological differences based on SES, parameterized by empirical studies.

#### Contact Network Model

In a contact network model, nodes represent individuals, and edges represent epidemiologicallyrelevant interactions between individuals. The degree of a node is the number of edges, or contacts, of the node, and the degree distribution of a network is the frequency distribution of node degrees within the population. To generate realistic contact networks to evaluate epidemic outcomes, we used an egocentric exponential random graph model (ERGM) [49]. An egocentric ERGM allows for the construction of sociocentric networks based on egocentrically sampled data, in which participants (or *egos*) report the identity of their contacts (or *alters*), who may or may not be study participants. Our egocentric ERGM model was based on the POLYMOD dataset, a large, egocentric contact survey that took place across several countries in Europe to identify close interactions of over 7000 individuals across eight European countries [34].

Nodes in the network had the following attributes: (a) age, grouped as infants-toddlers (age 0-4), school-aged children (age 5-18), adults (age 19-64), and elderly (age 65-100); (b) sex, classified as male or female; (c) contact location, in which a node can have known home contacts and known school or work contacts; (d) education level as a proxy for socioeconomic status [33], grouped as low education (less than a high school education), medium education (high school or vocational school education), or high education (any university education or beyond). Age and sex were available in the data for egos and alters, while education level was only provided for egos. Therefore, it was assumed that an ego’s work contacts had the same education level based on their occupation, and that an ego’s home contacts had the same education level as an indicator of household socioeconomic status. To represent communities with different SES compositions, we resampled additional low education egos from the low education sample in the POLYMOD dataset. We produce networks composed of approximately 20-60% low education individuals (Table S3).

The model was fit using the ERGM package [50, 51]. The best model was selected based on collinearity criteria and goodness of fit to the POLYMOD data. From the best fit ERGM model, we simulated 5 networks. Additional model details, including model terms (Table S1), collinearity (Table S2), model diagnostics, and goodness of fit (Fig S5 - Fig S30) can be found in the Supplement. Random regular networks of the same size and mean degree were also generated as null networks to evaluate the effect of contact heterogeneity. We used the Networkx package for network generation and analysis [52].

#### SES-based Epidemiological Model

Chain binomial SEIR (Susceptible-Exposed-Infected-Recovered) simulations were performed on the networks generated by the egocentric ERGM model and the control networks to examine the spread of a respiratory infection, like influenza, through a naive population. Model parameters pertinent to seasonal influenza spread were selected from literature (Table S4) [53, 54].

Five hypothesized drivers for increased influenza in low SES populations were integrated into the epidemiological simulations. Each hypothesized driver represents a health behavior or physiological factor, and is represented by a single parameter, the value of which was selected from pertinent literature (Table S4). Social contact differences represents the SES-based social contact rates of individuals, and thus is represented by the ERGM-generated networks. The remaining factors are:

- Low vaccine uptake: Individuals may be vaccinated before the start of the season with a perfectly efficacious vaccine. Vaccinated nodes were randomly selected and removed from the network. Vaccination coverage is parameterized by *δ* and *δ*_*low*_ in high- and low-SES individuals, respectively. The value of delta was based on a US population survey of vaccine coverage related to education level [19].
- High susceptibility: Those who experience a more stressful environment are more susceptible to infection, and thus have a greater probability of becoming infected upon contact with an infected individual. Susceptibility is parameterized by (*β* and (*β*_*low*_ in high- and low-SES individuals, respectively. This is based on an immune challenge experiment that found that those of high SES were about half as likely to become infected with a cold compared to those of low SES [22].
- Low healthcare utilization: Infected individuals who do not seek healthcare and receive antivirals have a longer infectious period, based on a model of within-host and population-level dynamics [55]. The proportion of the infected population seeking healthcare is parameterized by *γ* and *γ*_*low*_ in high- and low-SES individuals, respectively.

Low sickness absenteeism: Infected individuals may exhibit sickness absenteeism from school or work if they have access to leave and care at home. Those exhibiting sickness absenteeism remove school or work contacts. Access to sickness absenteeism is parameterized by ρ and ρ_*low*_ in high- and low-SES individuals, respectively. These values are based on rates of paid sick leave by education level in a survey across the US [56].

For our experimental design, each SES-based factor was tested separately and together on each network. Disease outbreaks for each treatment were simulated 200 times on each network, with 5 replicate networks. We also considered two controls to compare our experimental results: a) a homogeneous control, in which each factor was randomly distributed across a random regular network; b) a heterogeneous control, in which each factor was randomly distributed across the ERGM-generated networks.

### Modelling Impact of Disease in Low-SES Populations

To achieve an inferential understanding, we (a) integrated the network model findings with empirical ILI data for an estimate of ILI burden among low-SES individuals; and (b) fitted a spatial Bayesian hierarchical model with population-level covariates to account for measurement biases and improve our estimate of low-SES ILI burden at the population-level.

#### Spatial Inferential Model

We used a Bayesian spatial hierarchical model to estimate latent ILI cases among low SES individuals, accounting for measurement biases and county-level factors associated with ILI in low SES populations. We modeled low-SES ILI (*Y*_*it*_) in county i in flu season t as:

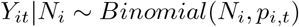

where *p*_*i,t*_ is the probability of detecting low-SES ILI cases, and *N*_*i*_ is the true ILI cases among low-SES individuals.

We modeled the probability of detection *p*_*i*_ as:

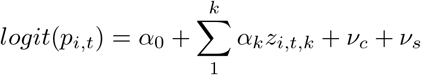

where *α*_*0*_ is the intercept, *α*_*k*_ represents the measurement process predictor variables, and *v*_*c*_ and *v*_*s*_ are group effects for county and state, respectively.

We modeled the latent low-SES ILI cases as:

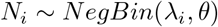

where the negative binomial distribution is parameterized by probability λ _*i*_ and size *θ*.

The λ _*i*_ is modeled by:

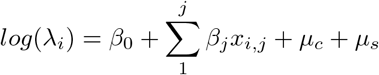

where (*β*_0_ is the intercept, (*β* _*j*_ represents coefficient estimates for low-SES ILI process covariates, and *μ*_*c*_ and *μ*_*s*_ represent county-level and state-level group effects, respectively. We performed approximate Bayesian inference using Integrated Nested Laplace Approximations (INLA) with the R-INLA package [57]. INLA has demonstrated computational efficiency for latent Gaussian models, produced similar estimates for fixed parameters as established implementations of Markov Chain Monte Carlo (MCMC) methods for Bayesian inference, and been applied to disease mapping and spatial ecology questions. We evaluated DIC, WAIC, model residuals and compared modeled and observed outcomes in order to assess model fit. Additional model details can be found in Fig S34, Fig S36, Fig S37.

#### Response data

We define the response in our model to be the observed influenza-like illness (ILI) burden in low-SES populations. In particular, we use influenza-like illness reports from a medical claims database from across the United States collected during 2002-2008. [Additional details on the dataset can be found in [31, 35]]. To normalize these observed counts, we divide ILI visits by visits for any diagnosis during the influenza season. These data are at the county-level but are not stratified by SES. This normalization was incorporated into the response data, rather than the measurement model, because the values vary greatly from season-to-season as database coverage increased, but INLA does not allow temporally varying measurement covariates [58]. To produce a county-level estimate of ILI in low-SES populations for our spatial model, we use the observed ILI burden in the total population and scale this by the proportion expected among low-SES individuals as predicted by the epidemiological model from the first part of our study (as summarized in Fig 2B). These scaled-up incidence rates were then multiplied by 1000 to be appropriately scaled for the model.

#### Covariate data

All covariate data are at the county level, and are centered and standardized. Covariate data cannot be included in a temporally-specific way in R-INLA, thus we assume that all values are constant over time from 2002-2008. We make the assumption that county characteristics remain relatively constant over time, and harness together covariate data from different years based on availability and coverage. All covariate data was evaluated for collinearity, and all included covariates had a VIF < 2. First, covariate data was included for the measurement submodel to characterize database coverage and population size. For database coverage, we used the number of physicians reporting the medical claims database, which was reported by the database and averaged over reported years. Additionally, the population of low SES individuals was included since that measures the size of the considered population. Low SES population size was measured as the county population size, reported by the US Census Bureau [59], multiplied by the percent of the population with less than a high school education from County Health Rankings [60]. Then, for the process model, covariate data were included as a marker for each hypothesized driver of low SES influenza. We ensured that all process covariate data pertained just to low SES populations. For a measure of susceptibility, reports of poor health in individuals with less than a high school education, divided by the sample size of low education individuals, were collected from the the Behavioral Risk Factor Surveillance System (BRFSS) from the CDC, which is available at the individual level and reported by county in 2012 [61]. For a measure of social cohesion, mean household size reported by those with less than a high school education was also collected from BRFSS. To measure access to healthcare, rates of reporting having health insurance, reporting having a personal doctor, and reporting avoiding healthcare due to cost by those of low education were divided by low education sample size from BRFSS. To measure sickness absenteeism, the rate of chronic sickness absenteeism, or students absent for more than 10 days, was collected from the US Department of Education [62]. This data was only available stratified by race, thus the chronic sickness absenteeism reports of Black students, divided by the number of Black students, was used due to the correlation between race and socioeconomic status in the US [63]. To measure vaccination, reports of adult vaccination in low education individuals were divided by the low education sample size in BRFSS [61]. Much of the data was available through BRFSS, which lacked coverage in many counties. Thus, counties with a low education sample size of less than 10 were omitted. Additionally, due to this sparse coverage in covariate data, we restricted our analyses to states that had complete covariate data for more than 50% of counties. This is to ensure that sparse covariate data does not skew the model, since we only want to provide inference for states that have enough data to provide reliable estimates. These challenges highlight the need for more high resolution data on low SES populations across the country. See supplement table for additional covariate data details (Table S5).

#### Imputation and Validation

Based on the assumption that counties that are close to one another are similar to one another, we imputed covariate values for missing counties in states that were included in the model. The model was run with only the counties that had complete covariate data. Then, for each missing county, we took the mean of the adjacent counties for each covariate value, to assign covariate values to the missing counties. We then used these imputed covariate values to calculate model estimates for the missing counties. The model estimates prior to imputation are available in Fig S35. We grouped the resulting full model estimates by county-level percent living in poverty, according to Small Area Income and Poverty Estimates reported by the US Census Bureau [64]. We collected incidence/incidence rate values reported by the same poverty level groupings from [12, 11]. Each set of incidence values was min-max normalized for comparison due to variations between reported value and population considered.

## Data Availability

All data associated with this study are provided in a GitHub repository: https://github.com/bansallab/fluSES

https://www.countyhealthrankings.org/

https://www.cdc.gov/brfss/index.html

https://www.ncdc.noaa.gov/cdo-web/

https://www2.ed.gov/datastory/chronicabsenteeism.html

https://github.com/bansallab/fluSES

## Acknowledgments

Research reported in this publication was supported by the National Institute Of General Medical Sciences of the National Institutes of Health under Award Number R01GM123007. The content is solely the responsibility of the authors and does not necessarily represent the official views of the National Institutes of Health. We also acknowledge support from the PhRMA Foundation and the Chateaubriand Fellowship Program. The funders had no role in study design, data collection and analysis, decision to publish, or preparation of the manuscript. We thank Håvard Rue for his development of and assistance with the R-INLA package.

## References

[1] Penman-Aguilar A, Talih M, Huang D, Moonesinghe R, Bouye K, Beckles G. Measurement of Health Disparities, Health Inequities, and Social Determinants of Health to Support the Advancement of Health Equity. J Public Helath Manag Pract. 2016;22(Suppl1):S33–S42.

[2] Adler NE, Newman K. Socioeconomic disparities in health: Pathways and policies. Health Affairs. 2002;21(2):60–76.

[3] Murray CJ, Kulkarni SC, Michaud C, Tomijima N, Bulzacchelli MT, Iandiorio TJ, et al. Eight Americas: investigating mortality disparities across races, counties, and race-counties in the United States. PLoS medicine. 2006;3(9).

[4] Bosworth B. Increasing Disparities in Mortality by Socioeconomic Status. Annual Review of Public Health. 2018;39(1):237–251.

[5] Centers for Disease Control and Prevention. Influenza; 2019.

[6] Biggerstaff M, Jhung MA, Reed C, Garg S, Balluz L, Fry AM, et al. Impact of medical and behavioural factors on influenza-like Illness, healthcare-seeking, and antiviral treatment during the 2009 H1N1 pandemic — United States, 2009–2010. Epidemiol Infect. 2014;142(1):114–125.

[7] Lowcock EC, Rosella LC, Foisy J, McGeer A, Crowcroft N. The social determinants of health and pandemic h1n1 2009 influenza severity. American Journal of Public Health. 2012;102(8):51–58.

[8] Rutter PD, Mytton OT, Mak M, Donaldson LJ. Socio-economic disparities in mortality due to pandemic influenza in England. International Journal of Public Health. 2012;57(4):745–750.

[9] Galvin JR, Cartter ML, Sosa L. Neighborhood Socioeconomic Status Among Children Hospitalized With Influenza: New Haven County, Connecticut, 2003-2010. Connecticut Epidemiologist. 2010;30(6):21–24.

[10] Yousey-Hindes KM, Hadler JL. Neighborhood socioeconomic status and influenza hospitalizations among children: New Haven County, Connecticut, 2003-2010. American Journal of Public Health. 2011;101(9):1785–1789.

[11] Tam K, Yousey-hindes K, Hadler L. Influenza-related hospitalization of adults associated with low census tract socioeconomic status and female sex in New Haven County, Connecticut, 2007-2011. Influenza and other Respiratory Viruses. 2014;8(3):274–281.

[12] Hadler JL, Yousey-hindes K, Pérez A, Anderson EJ, Bargsten M. Influenza-Related Hospitalizations and Poverty Levels — United States, 2010 – 2012. CDC Morbidity and Mortality Weekly Report. 2016;65(5):101–105.

[13] Crighton EJ, Elliott SJ, Moineddin R, Kanaroglou P, Upshur R. A spatial analysis of the determinants of pneumonia and influenza hospitalizations in Ontario (1992-2001). Social Science and Medicine. 2007;64(8):1636–1650.

[14] Grantz KH, Rane MS, Salje H, Glass GE, Schachterle SE. Disparities in influenza mortality and transmission related to sociodemographic factors within Chicago in the pandemic of 1918. PNAS. 2016;.

[15] Murray CJ, Lopez AD, Chin B, Feehan D, Hill KH. Estimation of potential global pandemic influenza mortality on the basis of vital registry data from the 1918–20 pandemic: a quantitative analysis. The Lancet. 2006;368(9554):2211–2218.

[16] Mamelund SE. 1918 pandemic morbidity: The first wave hits the poor, the second wave hits the rich. Influenza and other respiratory viruses. 2018;12(3):307–313.

[17] Cordoba E, Aiello AE. Social Determinants of Influenza Illness and Outbreaks in the United States. NCMJ. 2016;77(5):341–345.

[18] Charland KM, Brownstein JS, Verma A, Brien S, Buckeridge DL. Socio-economic disparities in the burden of seasonal influenza: The effect of social and material deprivation on rates of influenza infection. PLoS ONE. 2011;6(2):1–5.

[19] Linn ST, Guralnik JM, Patel KK. Disparities in Influenza Vaccine Coverage in the United States, 2008. J Am Geriatric Soc. 2010;58(7):1333–1340.

[20] Derigne L, Stoddard-dare P, Quinn L. Workers without Paid Sick Leave less Likely to Take Time Off For Illness or Injury Compared to those with Paid Sick Leave. Health Affairs. 2016;35(3):520–527.

[21] Cohen F, Kemeny ME, Zegans LS, Johnson P, Kearney KA, Stites DP. Immune function declines with unemployment and recovers after stressor termination. Psychosomatic Medicine. 2007;69(3):225–234.

[22] Cohen S, Adler N, Alper CM, Doyle WJ, Treanor JJ, Turner RB. Objective and Subjective Socioeconomic Status and Susceptibility to the Common Cold. Health Psychology. 2008;27(2):268–274.

[23] Berendes D, Andujar A, Barrios LC, Hill V. Associations Among School Absenteeism, Gastrointestinal and Respiratory Illness, and Income — United States, 2010 – 2016. CDC Morbidity and Mortality Weekly Report. 2019;68(9).

[24] Blendon RJ, Koonin LM, Benson JM, Cetron MS, Pollard WE, Mitchell EW, et al. Public Response to Community Mitigation Measures for Pandemic Influenza. Emerging Infectious Diseases. 2008;14(5).

[25] Cardoso MRA, Cousens SN, D. Góes Siqueira Lf, Alves FM, D’Angelo LAV. Crowding: Risk factor or protective factor for lower respiratory disease in young children? BMC Public Health. 2004;4:1–8.

[26] Sloan C, Chandrasekhar R, Mitchel E, Schaffner W, Lindegren ML. Socioeconomic Disparities and Influenza Hospitalizations, Tennessee, USA. Emerging Infectious Diseases. 2015;21(9).

[27] Munday JD, Van Hoek AJ, Edmunds WJ, Atkins KE. Quantifying the impact of social groups and vaccination on inequalities in infectious diseases using a mathematical model. BMC Medicine. 2018;16(1):1–12.

[28] Hyder A, Leung B. Social deprivation and burden of influenza: Testing hypotheses and gaining insights from a simulation model for the spread of influenza. Epidemics. 2015;11:71–79. Available from: http://dx.doi.org/10.1016/j.epidem.2015.03.004.

[29] Kumar S, Piper K, Galloway DD, Hadler JL, Grefenstette JJ. Is population structure suffcient to generate area-level inequalities in influenza rates? An examination using agent-based models. BMC Public Health. 2015;15(1):1–12.

[30] World Health Organization. WHO global technical consultation: global standards and tools for influenza surveillance (WHO/HSE/GIP/2011.1). 2011;(March). Available from: http://whqlibdoc.who.int/hq/2011/WHO_HSE_GIP_2011.1_eng.pdf?ua=1.

[31] Lee EC, Arab A, Goldlust SM, Grenfell B. Deploying digital health data to optimize influenza surveillance at national and local scales. PLoS Computational Biology. 2018;14(3):1–23.

[32] Scarpino SV, Scott JG, Eggo RM, Clements B, Dimitrov NB, Meyers LA. Socioeconomic bias in influenza surveillance. PLoS Computational Biology. 2020;16(7). Available from: http://arxiv.org/abs/1804.00327.

[33] Thomson S. Achievement at school and socioeconomic background—an educational perspective. npj Science of Learning. 2018;3(1):5. Available from: http://www.nature.com/articles/s41539-018-0022-0.

[34] Mossong J, Hens N, Jit M, Beutels P, Auranen K, Mikolajczyk R, et al. Social contacts and mixing patterns relevant to the spread of infectious diseases. PLoS Medicine. 2008;5(3):0381–0391.

[35] Viboud C, Charu V, Olson D, Ballesteros S, Gog J, Khan F, et al. Demonstrating the use of high-volume electronic medical claims data to monitor local and regional influenza activity in the US. PLoS ONE. 2014;9(7).

[36] Nettelman MD, White T, Lavoie S, Chafin C. School Absenteeism, Parental Work Loss, and Acceptance of Childhood Influenza Vaccination. The American Journal of Medical Sciences. 2001;321(3):178–180.

[37] Neuzil KM, Hohlbein C, Zhu Y. Illness Among Schoolchildren During Influenza Season. Archives of Pediatrics & Adolescent Medicine. 2002;156(10):986.

[38] Clemans-Cope L, Perry CD, Kenney GM, Pelletier JE, Pantell MS. Access to and use of paid sick leave among low-income families with children. Pediatrics. 2008;122(2).

[39] Aronsson, G, Gustafsson, K, & Dallner M. Sick but yet at work. An empirical study of sickness presenteeism. Journal of Epidemiology and Community Health. 2000;54:502–509.

[40] Freimuth VS, Jamison A, Hancock G, Musa D, Hilyard K, Quinn SC. The Role of Risk Perception in Flu Vaccine Behavior among African-American and White Adults in the United States. Risk Analysis. 2017;37(11):2150–2163.

[41] Lucyk K, Simmonds KA, Lorenzetti DL, Drews SJ, Svenson LW, Russell ML. The association between influenza vaccination and socioeconomic status in high income countries varies by the measure used: a systematic review. BMC Medical Research Methodology. 2019;19(1).

[42] Endrich MM, Blank PR, Szucsb TD. Influenza vaccination uptake and socioeconomic determinants in 11 European countries. Vaccine. 2009;27(30):4018–4024.

[43] Bansal S, Grenfell BT, Meyers LA. When individual behaviour matters: Homogeneous and network models in epidemiology. Journal of the Royal Society Interface. 2007;4(16):879–891.

[44] Cauchemez S, Bhattarai A, Marchbanks TL, Fagan RP, Ostroff S, Ferguson NM, et al. Role of social networks in shaping disease transmission during a community outbreak of 2009 H1N1 pandemic influenza. Proceedings of the National Academy of Sciences. 2011;108(7):2825–2830. Available from: http://www.pnas.org/cgi/doi/10.1073/pnas.1008895108.

[45] Kennedy BP, Kawachi I, Glass R, Prothrow-Stith D. Income distribution, socioeconomic status, and self rated health in US : multilevel analysis. British Medical Journal. 1999;318(7195):1417–1418.

[46] Noel R. Race, Economics, and Social Status. 2018;(May):2–9.

[47] Raifman MA, Raifman JR. Disparities in the Population at Risk of Severe Illness from COVID-19 by Race/Ethnicity and Income. American Journal of Preventative Medicine. 2020;59(1):137–139.

[48] Zipfel, C and Bansal, S. Bansal Lab Github Account; 2020. Available from: https://github.com/bansallab/fluSES.

[49] Krivitsky PN, Handcock MS, Morris M. Adjusting for network size and composition effects in exponential-family random graph models. Statistical Methodology. 2011;8(4):319–339. Available from: http://dx.doi.org/10.1016/j.stamet.2011.01.005.

[50] Handcock MS, Hunter DR, Butts CT, Goodreau SM, Krivitsky PN, Morris M. ergm: Fit, Simulate and Diagnose Exponential-Family Models for Networks; 2019.

[51] Hunter DR, Handcock MS, Butts CT, Goodreau SM, Morris M. ergm: A Package to Fit, Simulate and Diagnose Exponential-Family Models for Networks. Journal of Statistical Software. 2008;24(3):1–29.

[52] Hagberg AA, Schult DA, Swart PJ. Exploring network structure, dynamics, and function using NetworkX. Proceedings of the 7th Python in Science Conference (SciPy2008). 2008;Gäel Varoq:11–15.

[53] Biggerstaff M, Jhung MA, Reed C, Fry AM, Balluz L, Finelli L. Influenza-like illness, the time to seek healthcare, and influenza antiviral receipt during the 2010–11 influenza season — United States. J Infect Dis. 2014;210(4):535–544.

[54] Carrat F, Vergu E, Ferguson NM, Lemaitre M, Cauchemez S, Leach S, et al. Timelines of infection and disease in human influenza: A review of volunteer challenge studies. American Journal of Epidemiology. 2008;167(7):775–785.

[55] Pepin KM, Riley S, Grenfell BT. Effects of Influenza Antivirals on Individual and Population Immunity Over Many Epidemic Waves. Epidemiol Infect. 2013;141(2):366–376.

[56] Piper K, Youk A, James AE, Kumar S. Paid sick days and stay-At-home behavior for influenza. PLoS ONE. 2017;12(2):1–13.

[57] Rue H, Martino S, Chopin N. Approximate Bayesian inference for latent Gaussian models using integrated nested Laplace approximations (with discussion). Journal of the Royal Statistical Society, Series B. 2009;71(2):319–392. Available from: www.r-inla.org.

[58] Meehan TD, Michel NL, Rue H. Estimating animal abundance with N-mixture models using the R-INLA package for R. 2017;(May). Available from: http://arxiv.org/abs/1705.01581.

[59] Bureau UC. County Population Totals: 2010-2019. 2020;.

[60] University of Wisconsion Population Health Institute. County Health Rankings and Roadmaps; 2019.

[61] Centers for Disease Control and Prevention. Behavioral Risk Factor Surveillance System; 2012.

[62] US Department of Education. Chronic Absenteeism in the Nation’s Schools. 2019;Available from: https://www2.ed.gov/datastory/chronicabsenteeism.html.

[63] LaVeist TA. Disentangling race and socioeconomic status: A key to understanding health inequalities. Journal of Urban Health. 2005;82(SUPPL. 3).

[64] U S Census Bureau. Small Area Income and Poverty Estimates (SAIPE) Program; 2020. Available from: https://www.census.gov/programs-surveys/saipe.html.

